# Prediction of severe COVID-19 cases requiring intensive care in Osaka, Japan

**DOI:** 10.1101/2021.06.05.21258407

**Authors:** Junko Kurita, Tamie Sugawara, Yasushi Ohkusa

**Affiliations:** Department of Nursing, Tokiwa University, Ibaraki, Japan; National Institute of Infectious Diseases, Tokyo, Japan

**Keywords:** COVID-19, medical exhaustion, prediction, severe case, intensive care unit

## Abstract

**Background:** To avoid exhaustion of medical resources by COVID-19 care, policy-makers must predict care needs, specifically estimating the proportion of severe cases likely to require intensive care. In Osaka prefecture, Japan, the number of these severe cases exceeded the capacity of ICU units prepared for COVID-19 from mid-April, 2021.

**Objective:** This study used a statistical model to elucidate dynamics of severe cases in Osaka and validated the model through prospective testing.

**Methods:** The study extended from April 3, 2020 through April 26, 2021 in Osaka prefecture, Japan prefecture. We regressed the number of severe cases on the number of severe cases the day prior and the newly onset patients of more than 21 days prior.

**Results:** We selected the number of severe cases the day prior and the number of newly onset patients on 21 and 28 days prior as explanatory variables for explaining the number of severe cases based on the adjusted determinant coefficient. The adjusted coefficient of determination was greater than 0.995 and indicated good fit. Prospective out of sample three-week prediction forecast the peak date precisely, but the level was not t.

**Discussion and Conclusion:** A reason for the gap in the prospective prediction might be the emergence of variant strains.

## Introduction

Since mid-April, the number of severe cases in Osaka prefecture, Japan, with its 8.8 million residents, exceeded the intensive care unit (ICU) capacity prepared for COVID-19 patients. As shown in Figure 1, the maximum capacity was 365 beds on May 5 and 6, 2021. Prospective estimation of the number of severe cases is necessary to plan and activate countermeasures. Actually, at least one researcher used a simple exponentially increasing figure to predict 500 severe cases by the end of April on April 14 [1]. Although its validity was not evaluated, 500 severe cases represent more than twice the capacity for it. Therefore, the present study examines provision of the prediction model and its validity.

**Figure 1:**
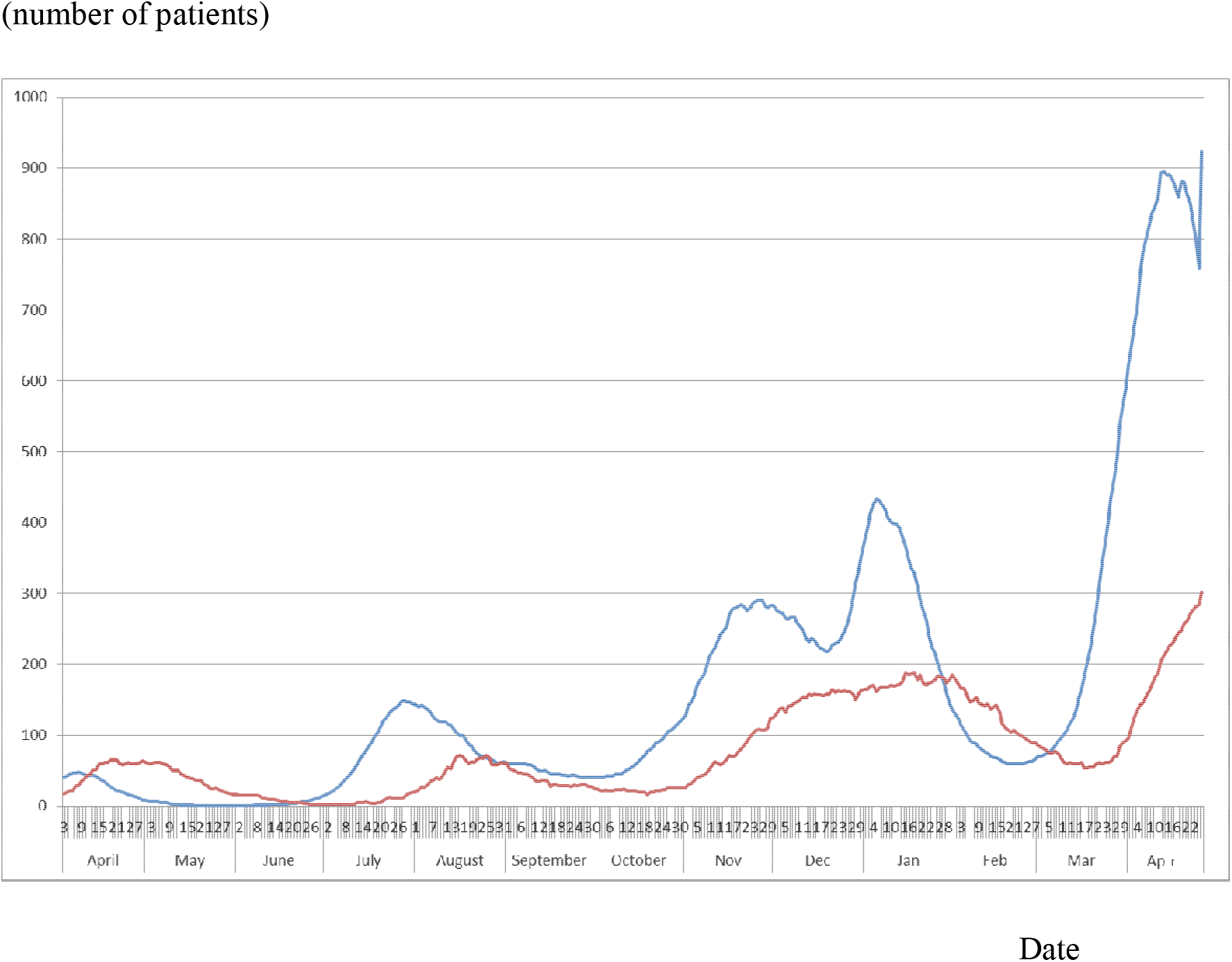
Epidemic curve of the symptomatic confirmed cases and number of severe cases in Osaka, April 3, 2020 – April 26, 2021. Date Notes: The blue line represents the epidemic curve of the symptomatic confirmed cases on the onset date. The red line represents the number of severe cases.

## Methods

Severe cases of COVID-19 were defined as requiring ICU admission. The study period extended from April 3, 2020 to April 26, 2021. Out of sample prediction was performed for three weeks of April 27 through May 17, 2021. The study area was Osaka prefecture in Japan.

We regressed the number of severe cases on the lagged number of severe cases and the newly onset patients reported more than 21 days prior. Then we selected a formula maximizing the adjusted coefficient of determination.

The study area was Osaka prefecture. The daily newly confirmed cases and information about severe cases in Osaka were published by the Osaka prefectural government [2]. Epidemic curves, which are newly onset patients by onset date, were calculated through empirical distribution of the incubation period and the reporting delay from onset to reporting, similarly to earlier studies [3, 4].

Finally, to evaluate the model’s predictive power, we performed out of sample forecasting prospectively for three weeks from April 26, 2021 until May 17, 2021, fixing the parameters as estimated. Evaluation measures for the precision of prediction were correlation coefficients and discrepancy rates, which were defined as the percentage of the average of the absolute difference between the observed and the predicted, divided by the observed, weighted by the observed [6]. We adopted 5% as the significance level.

## Ethical considerations

All information used for this study was published by the Osaka prefectural government [2]. Therefore no ethical issue was posed by the procedures used to conduct this study.

## Results

In Osaka, we observed 76,483 COVID-19 patients severe cases and 29,894 severe cases day up through April 26, 2021. Figure 1 depicts severe cases and newly onset patients each day. Severe cases fluctuate with a lag after the number of onset patients. Table 1 presents the estimation results. Based on adjusted coefficients of determination, we selected the model with the number of severe cases the day prior and the number of newly onset patients 21 and 28 days prior. Its adjusted coefficient of determination was higher than 0.995. Figure 2 depicts the observed and fitted numbers of severe cases. It is apparent that these were almost mutually equal.

**Table 1:** Estimation results for the number of severe cases

| Explanatory variable | Estimated coefficient | *p*-value | Estimated coefficient | *p*-value | Estimated coefficient | *p*-value | Estimated coefficient | *p*-value | Estimated coefficient | *p*-value |
| --- | --- | --- | --- | --- | --- | --- | --- | --- | --- | --- |
| constant | 16.4607 | .000 | 0.1430 | .646 | 0.1783 | .564 | -0.19874 | .568 | 0.1903 | .544 |
| Lag of the number of severe cases |  |  |  |  |  |  |  |  |  |  |
| *t*-1 |  |  | 1.0185 | .000 | 1.1426 | .000 | 0.99658 | .000 | 1.0295 | .000 |
| *t*-2 |  |  |  |  | -0.1298 | .019 |  |  |  |  |
| Lag of the number of newly onset cases |  |  |  |  |  |  |  |  |  |  |
| *t*-21 | 0.4079 | .000 | 0.0319 | .000 | 0.0292 | .000 | 0.00859 | .070 | 0.0196 | .040 |
| *t*-28 | 0.0686 | .005 | -0.0427 | .000 | -0.0370 | .000 |  |  | -0.0249 | .068 |
| *t*-35 |  |  |  |  |  |  |  |  | -0.0123 | .163 |
| Number of observations | 361 |  | 361 |  | 361 |  | 368 |  | 354 |  |
| Adjusted *R*<sup>2</sup> | 0.8957 |  | 0.9971 |  | 0.9971 |  | 0.9962 |  | 354 |  |

Note: Rows with “*t*-1” or “*t*-2” show the estimated coefficients and *p*-values for the
number of severe cases yesterday or the day before yesterday. Row with “*t*-21,” “*t*-28”,
and “*t*-35” respectively show the estimated coefficients and *p*-values for the number of
newly onset patients 21, 28, and 35 days prior. Based on adjusted coefficients of
determination, we selected the model with one day lag of the number of severe case and
the number of newly onset patients at 21 and 28 days prior.

**Figure 2:**
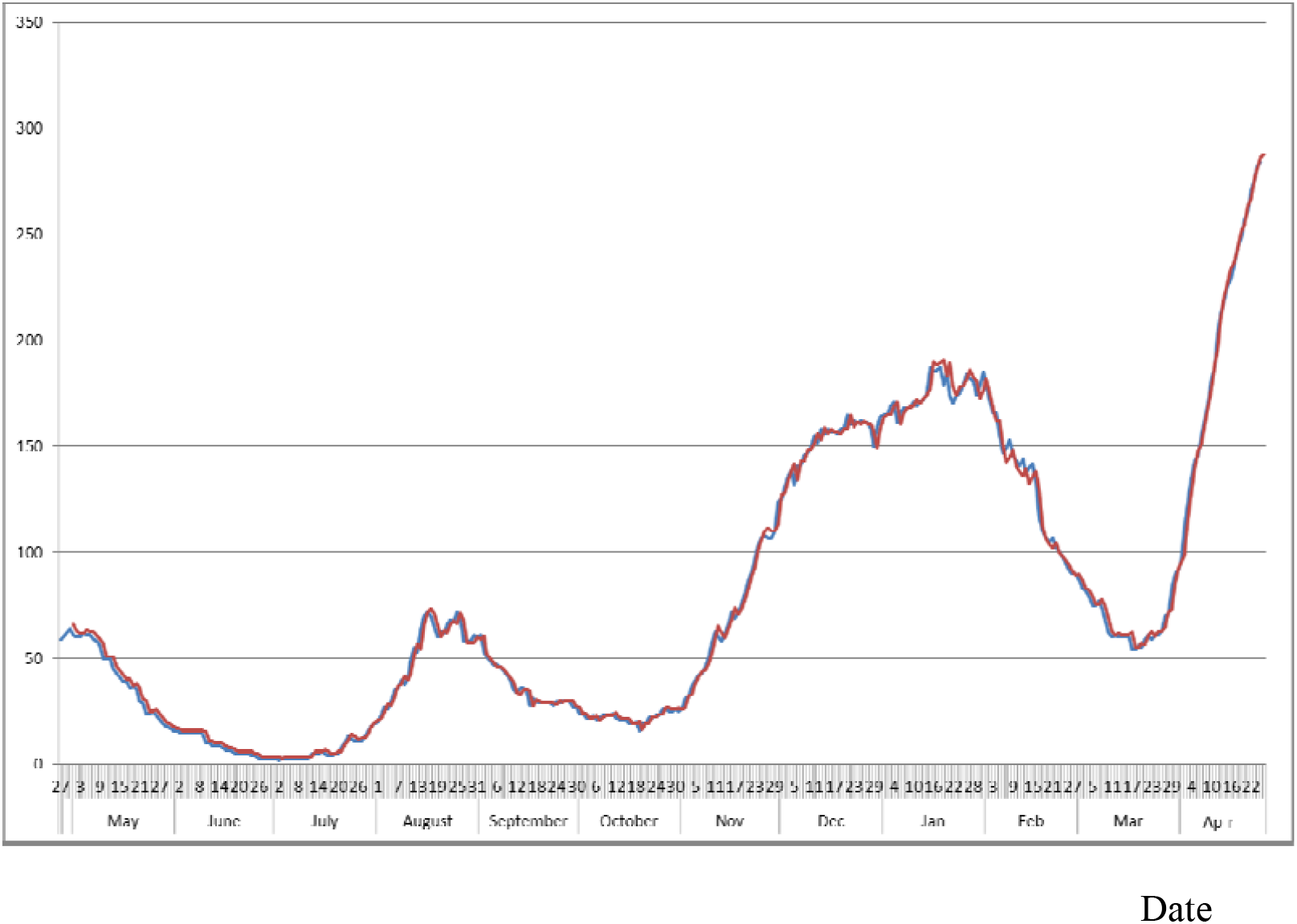
Estimated and observed number of severe cases in Osaka, April 3, 2020 – April 26, 2021. (number of patients) Note: The blue line shows the observed number of severe cases. The red line represents the estimated number of severe cases.

Figure 3 depicts out of sample three-week forecasting prospectively for 21 days from April 26, 2021. The observed peak was May 4, 2021, but the peak in forecasting was April 30, 2021. Although there were some gaps between the two dates, because the observed and predicted values were almost flat for a few days before and after, the model can predict the peak date about one week prior. Nevertheless, the peak was not predicted precisely. Prediction was far less than the observed value as time progressed.

**Figure 3:**
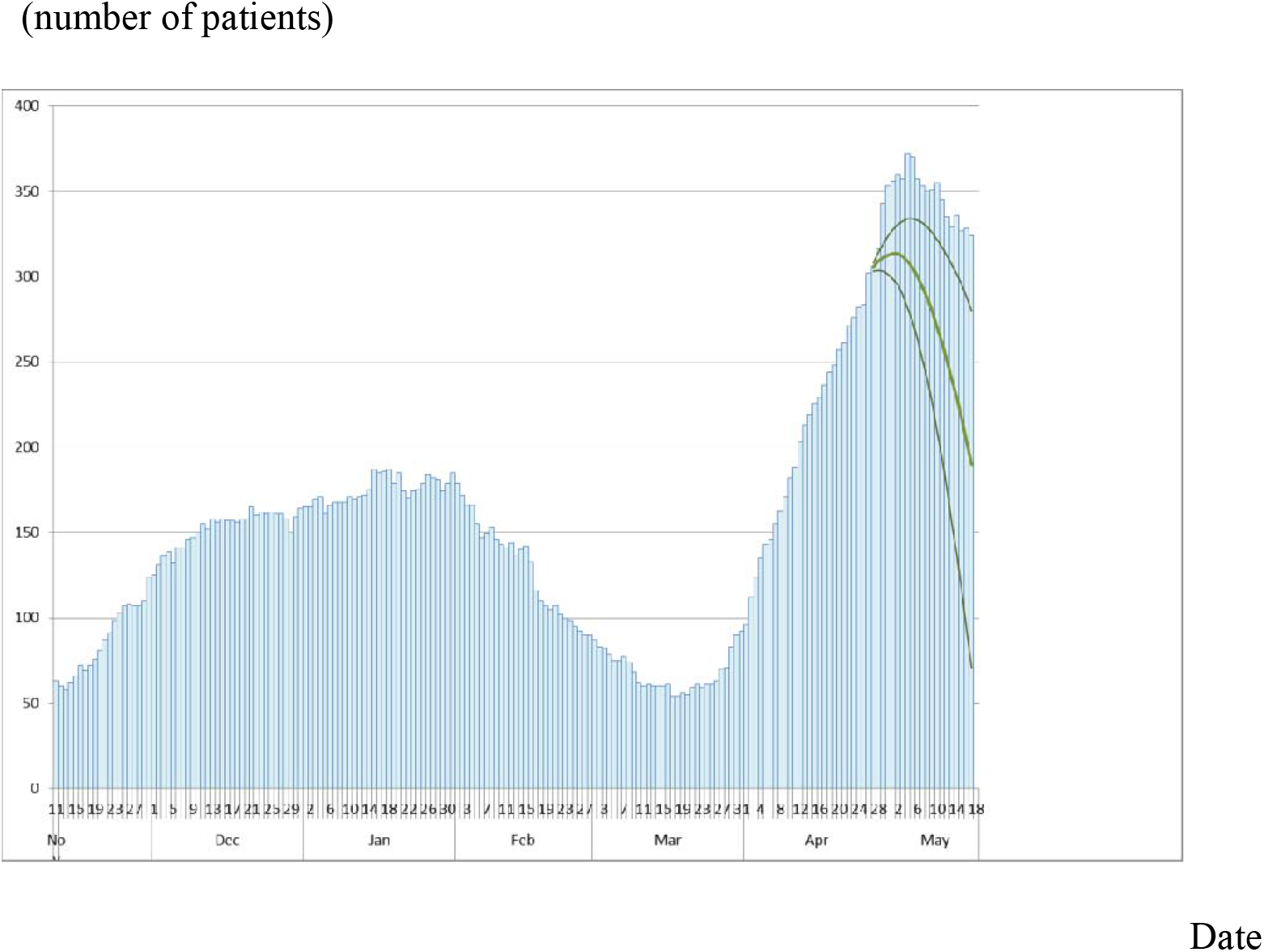
Observed data of severe cases and its forecasting as of April 26, 2021. Notes: Bars represent data of severe cases observed daily in Osaka from December 3, 2020. The bold line represents a forecast based on estimation results presented in Table 1. Thin lines represent its 95% CI.

Actually, correlation coefficients were 0.494, which was inferred as significant. The discrepancy rate was 19.52%. If the length of prediction is shortened from three weeks, then these numbers were −0.063 and 13.8%. These were 0.965 and 8.95%.

## Discussion

Many mathematical models have been applied to the outbreak of COVID-19 worldwide [3–5]. Unfortunately, these have not succeeded in explaining the course of COVID-19 outbreaks with several peaks and fluctuations. Even in Osaka, estimation based on a mathematical model yielded overestimations by more than 40% [1]. Especially in Japan, four peaks were experienced through the end of April, as shown in Figure 1. The first peak was that of earlier April. The second one was at the end of July. The first peak might explain the mathematical model with restrictions against excursions from home [6] because, since mid-March, the rate of going out declined, as shown in Apple data. However, the second and additional peaks were inexplicable if using the simple mathematical model. Herd immunity, of course, was not achieved. No lock-down was activated in Japan at all. Probably, the “new normal” lifestyle with mask-wearing, social distancing, and restrictions against numerous dining or night-life establishments became common.

Actually, from late July to the end of December, the government encouraged domestic travel. It promoted the spread of the outbreak. However, the outbreak was contained in August. Therefore, to date, no mathematical model has been sufficient to explain the outbreak in Japan. The policy for severe cases cannot be discussed using mathematical models.

Instead, we used a statistical model. It might always show some association of variables for which there is no causality. We showed clear association among severe cases and patients who had experienced onset 21 days prior.

We used no information before *t*-21. Reporting was usually delayed from onset up to 31 days [3, 4]. Therefore, the number of newly onset patients at the same day was that which was usually developed dynamically for at least two weeks. Because information of the final two weeks before the estimate is unreliable, we ignored information during the final two weeks of the study period. Moreover, because the epidemic curve changes gradually, information related to day *t* is correlated closely with information related to day *t*-1. Therefore, we used no information related to consecutive days. We used the number of newly onset patients with 21 and 28 days prior as explanatory variables.

Prospective out of sample forecasting in Figure 3 shows good fit. Especially, the peak date was predicted precisely. However, the number of cases at that peak cannot not be predicted precisely. Correlation coefficients for three week prediction were not high. The peak was found to be the second week. Therefore, correlation for the two-week prediction was negative and not significant. The discrepancy rate was also not small. Even for the first week, it was only about 9%. Therefore, we cannot evaluate that the model is able to predict the level of severe cases.

A potential reason might be the variant strain. It emerged in March, 2021 and spread in April [8]. Even though the study period included these days, the model mainly reflected the original strain comparison with the variant strain. Therefore, the gap within which the model cannot be predicted might be caused by higher pathogenies in the variant strain than in the original strain.

Some limitations might have affected this study. First, we used dummy variables after June to ascertain the disease’s decreasing severity. However, we do not understand the reason why that decreasing severity occurred at that time. That diminished severity might be attributable to COVID-19 virus mutation. Alternatively, treatment for patients might have improved since June. Younger and therefore milder patients accounted for a larger proportion of the patients than before June. Moreover, the disease severity can be expected to decrease if a new drug were developed. Alternatively, it might increase if some mutation were to strengthen its pathogenicity. Severity should be monitored to improve the fit of the statistical model.

Secondly, definitions of severe cases differed among prefectures, even in Japan. Moreover, they probably varied among countries. It remains unknown whether the approach used for the present study is applicable to prefectures other than Osaka or to other countries with other definitions of severity. The robustness of usefulness of the statistical model must be verified for other areas in Japan and other countries.

Thirdly, we might specifically examine only elderly patients or patients older than 50 as explanatory variables. Exploration of that point remains as a challenge for future research efforts.

Fourthly, Osaka has experienced higher severity than Tokyo. In actuality, on April 26, 2021, Osaka had 306 severe cases, with only 55 severe cases reported among Tokyo’s 13 million residents. Therefore, the proportion of severe cases per capita in Osaka was 8.2 times greater than in Tokyo. Of course, this proportion fluctuates daily, but this great difference must be investigated to elucidate Osaka situations of the severe cases. Difference in the prevalence of variant strain with mutation at N501Y or E484K between the two areas might lead to differences in severity. Alternatively, differences in the situations of underlying diseases or countermeasures might explain the difference. Resolving this difficulty is anticipated as our next challenge.

## Conclusion

Results demonstrated that the model can predict the peak date very well. However, quantitative prediction of the number of severely ill infected persons was much lower than the observed level. The present study is based on the authors’ opinions. Its results do not reflect any stance or policy of their professionally affiliated bodies.

## Data Availability

Osaka Prefectural Government. The Situation of Monitored Items in Osaka Model. (in Japanese)

http://www.pref.osaka.lg.jp/iryo/osakakansensho/corona_model.html

## Acknowledgments

We acknowledge the great efforts of all staff at public health centers, medical institutions, and other facilities who are fighting the spread and destruction associated with COVID-19. This study represents the authors’ opinion. It does not reflect any stance of our affiliation.

